# Stagnation in quality of next-generation sequencing assays for the diagnosis of hereditary hematopoietic malignancies

**DOI:** 10.1101/2022.06.14.22276069

**Authors:** Gregory W. Roloff, Reid Shaw, Timothy E. O’Connor, Michael W. Drazer

## Abstract

**Importance:** Hereditary hematopoietic malignancies (HHMs) are hereditary cancer syndromes that constitute at least 14% of all myeloid malignancies, but genetic assays used to diagnose HHMs have historically been of variable quality. Here, we demonstrate that HHM assays continue to have persistent shortcomings. These diagnostic gaps place patients with HHMs at high risk for missed diagnoses, missed opportunities for cancer screening, and donor-derived leukemias following stem cell transplant.

**Objective:** To determine if the quality of HHM diagnostic assays has improved since 2020, when our group first demonstrated that most HHM diagnostic tests were insufficient for the accurate diagnosis of these syndromes. We hypothesized that the number of genes tested on each HHM assay increased from 2020 to 2022, in keeping with a more comprehensive sequencing approach.

**Design, Setting, and Participants:** We analyzed assays from eight commercial laboratories to determine which HHM-related genes were sequenced by these assays. We compared these assays to panels from 2020 to determine trends in sequencing quality.

**Results:** The majority of HHM diagnostic assays did not change over time and are insensitive for the detection of the full spectrum of HHM-related mutations. The majority (75%) of HHM assays do not sequence *CHEK2*, the gene most frequently mutated in HHMs, and 25% of HHM assays do not sequence *DDX41*, the second most frequently mutated HHM gene.

**Conclusions:** The quality of HHM diagnostic assays has stagnated despite the discovery of novel HHM-related genes as well as prior work demonstrating heterogeneity in quality of HHM testing. The majority of commercially available HHM tests remain insufficient for the diagnosis of the full spectrum of HHM-related germline mutations.

**Key Points:** *Question:* How have diagnostic assays for hereditary hematopoietic malignancies (HHMs) changed since 2020, when most HHM diagnostic assays were inadequate for the accurate diagnosis of HHMs?

*Findings:* Most HHM assays have significant deficiencies in quality and do not sequence the most relevant HHM-related genes. No meaningful improvements in the quality of HHM diagnostic testing have occurred since 2020.

*Meaning:* The quality of HHM diagnostic testing must be improved to universally include the most common HHM-related germline mutations. This will reduce the risk for false negatives, donor derived leukemias, improve genetic counseling, and improve screening for other HHM-related malignancies.

## Introduction

Hereditary hematopoietic malignancies (HHMs) are hereditary blood cancer syndromes driven by germline mutations. All HHMs to date have followed Mendelian inheritance patterns.^1^ The scientific basis and clinical recognition of HHMs has increased with the rapid uptake of next-generation sequencing (NGS) in research and clinical settings. The process of confirming germline mutations in HHMs requires multiple diagnostic steps, as peripheral blood represents tumor tissue that carries both germline and somatic mutations. Although germline mutations can be incidentally detected via tumor-only sequencing panels,^2^ the gold standard for identifying a germline mutation in a patient with an active hematopoietic malignancy is sequencing DNA from cultured skin fibroblasts.^3^

The accurate diagnosis of an HHM also requires NGS panels that sequence the full spectrum of genes involved in HHMs and sequencing techniques that are capable of detecting the various types of variants that drive HHMs (ie, single nucleotide variants, insertions/deletions, and structural events such as copy number changes). The number of HHM-related genes, however, has rapidly increased as the genetic basis of HHMs continues to be elucidated.^4^ This requires HHM diagnostic assays to be updated frequently in response to advances in scientific knowledge. One problem in the HHM field, however, is the quality of HHM diagnostic assays is highly variable.^5^ Most HHM assays used in 2020, for example, did not sequence all HHM-related genes. These omissions increase the risk for false negative test results, which place HHM patients at risk for complications such as donor-derived leukemias (when stem cells from a matched related donor who unknowingly carries an HHM related mutation are provided to an affected recipient), missed opportunities for cancer screening (in HHMs that also carry an increased risk for solid tumors, such as Li Fraumeni syndrome), and reduce opportunities for cascade testing of family members who may benefit from genetic counseling and personalized cancer screening. Other HHM assays in 2020 accepted non-germline tissues, such as DNA from peripheral blood mononuclear cells, as a permissible tissue source. Sequencing non-germline tissue, particularly from patients with active hematopoietic malignancies, increases the risk for false positive HHM test results, as NGS assays may detect both somatic mutations in residual tumor tissue or clonal hematopoiesis-related mutations.^2,6-9^

It is unclear how the landscape of HHM diagnostic testing has changed since we performed our original analysis of commercial testing practices in 2020.^5^ We identified all commercial diagnostic companies that offered NGS panels intended for HHM diagnosis in January 2022 to address this knowledge gap. We analyzed the spectrum of HHM-related genes sequenced by each commercial panel and compared these genes to prior offerings from the same company. Here, we demonstrate that the quality of commercial HHM test offerings has largely stagnated since 2020, despite a growing recognition of these hereditary blood syndromes and prior work demonstrating the low quality of HHM diagnostic assays.

## Methods

We identified HHM assays offered by commercial laboratories that we analyzed in our previous report. We also queried the US National Center for Biotechnology Information’s Genetic Testing Registry to identify all newly offered HHM test offerings since our original analysis. The updated analysis was performed in January 2022, one year after the print publication and sixteen months after the e-publication of our prior study.^5^ We focused our analysis on NGS panels for the diagnosis hereditary myelodysplastic syndrome and hereditary leukemia.

We evaluated all genes on HHM-related panels via a binary matrix approach. We calculated the proportion of panels that sequenced each HHM gene of interest and queried the Online Mendelian Inheritance in Man catalog to confirm the association of each HHM-related gene and a relevant HHM phenotype (**Supplementary Table 1**). Our analysis also included HHM-related genes that were most commonly mutated in a recent series of unselected patients with acute myeloid leukemia (**Supplementary Table 1**).^10^ The most frequently sequenced genes across HHM panels for 2020 and 2022 are plotted in **Figure 1**, defining a core set of HHM genes that were analyzed by the majority of commercial assays. We also plotted the proportion of HHM panels that sequenced a gene of interest in 2020 and 2022 to identify trends in gene-specific sequencing during this time period (**Figure 2, Supplementary Figure 1**).

**Figure 1.**
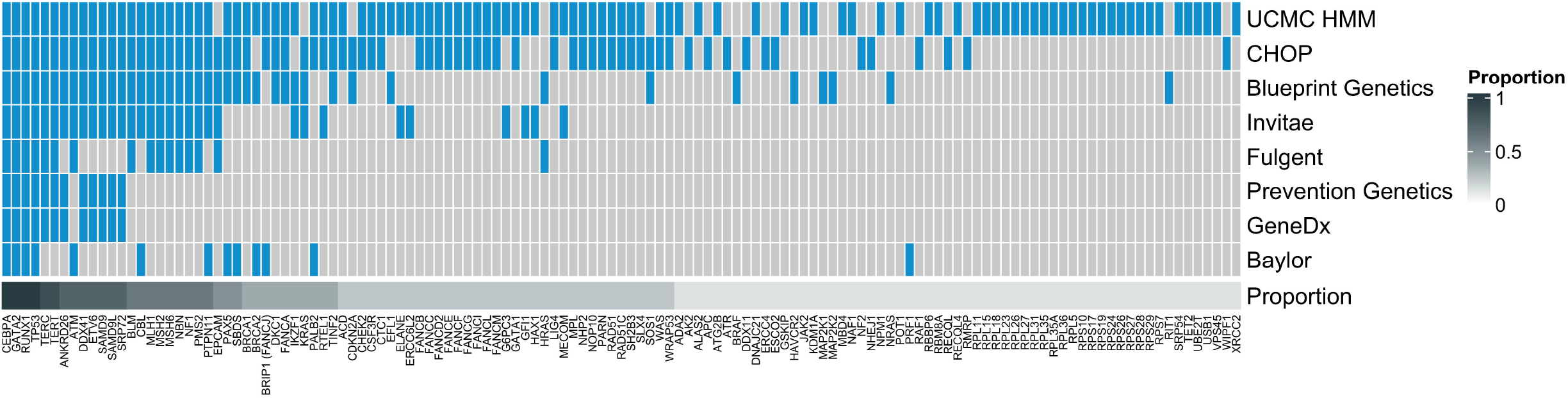
Comprehensive analysis of Hereditary Hematopoietic Malignancy diagnostic panels from eight commercial offerings. The heatmap depicts data via a binary matrix whereby any single gene included on one commercial panel is cross-referenced for inclusion on other assays analyzed. Genes are organized from left to right by decreasing proportion across all panels. Abbreviations used: UCMC HMM, University of Chicago Medical Center Hereditary Myeloid Malignancy Panel ; CHOP, Children’s Hospital of Philadelphia.

**Figure 2.**
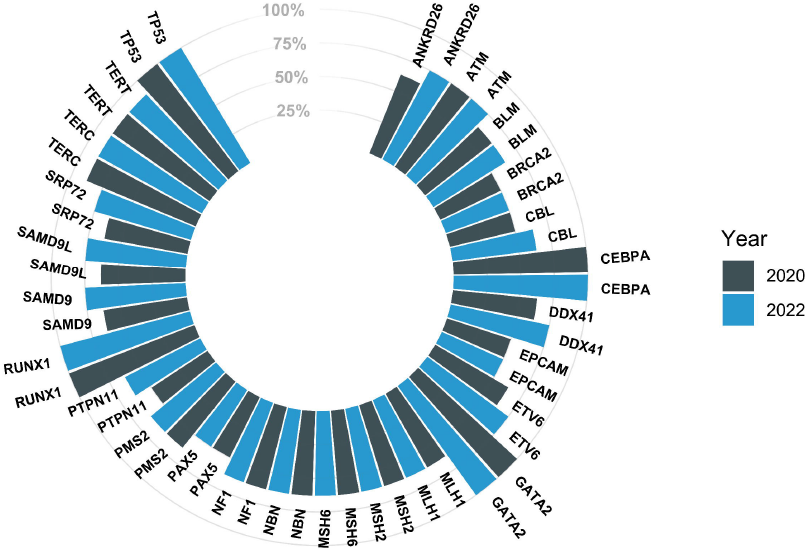
Analysis of commercial assay gene composition over time. This plot demonstrates percent composition of commercial panel inclusion for genes implicated in hereditary hematopoietic malignancies (HHMs). To emphasize high-yield HHM genes, only those included on at least half (4/8) of panels are displayed. Genes are depicted in pairs, demonstrating longitudinal change by the year analyzed.

## Results

We analyzed HHM panels from eight companies (**Figure 1**). Only four genes (*CEBPA, GATA2, RUNX1, TP53*) were sequenced by all eight panels. An additional 25 genes were sequenced on 50% or more of the panels. Only 25% of HHM panels sequenced *CHEK2*, the HHM gene most commonly mutated in Yang *et al*.,^10^ and only 75% of HHM panels sequenced *DDX41*, which was previously considered the gene that is most commonly mutated in HHMs.^11^ The discordance between the noted prevalence of mutations in HHM-associated genes in Yang *et al*. and their inclusion on commercial HHM panels is depicted in **Supplementary Figure 2**. Several important HHM genes, such as *BRCA1, BRCA2, CSF3R, FANCA*, and *MBD4* appeared on only a minority of panels (**Figure 1**). Furthermore *EPCAM*, which is not implicated in the etiology or pathogenesis of any known HHM, was present on half of all panels.

Trends in analysis of the 25 most-analyzed HHM-related genes from 2020 to January 2022 are shown in **Figure 2**. Most HHM assays that previously omitted critical HHM genes made no changes to their panels over this time period. This stagnancy persisted despite multiple associations between candidate genes and HHM phenotypes in public databases (**Supplementary Figure 2**). Only one laboratory made significant changes to their panel by adding eight HHM-related genes (*ANKRD26, CBL, DDX41, ETV6, PTPN11, SAMD9, SAMD9L, SRP72*; **Supplementary Figure 1**).

## Discussion

The accurate diagnosis of an HHM guides patient care in several clinically meaningful ways. First, the identification of an HHM mutation informs genetic counseling for the affected patient, while also streamlining cascade genetic testing for both affected and unaffected family members.^12^ Third, the identification of an HHM mutation refines the stem cell transplant process by allowing physicians to avoid using unaffected related mutation carriers as stem cell donors. This approach reduces the likelihood of donor-derived malignancies while also avoiding the currently unknown risks associated with the use of hematopoietic growth factors for stem cell mobilization in HHM mutation carriers.^13^ Finally, the identification of an HHM-related mutation may also guide therapeutic decision making. Recent evidence, for example, supports the utilization of lenalidomide for patients with myeloid malignancies driven by germline *DDX41* mutations even in the absence of the recurrent del(5q) abnormality.^14,15^

Here, we provide further evidence that the majority of HHM diagnostic assays remain inadequate for the accurate of diagnosis of these syndromes. Stagnation in the quality of HHM diagnostic assays has occurred despite the discovery of novel HHM-related genes, as well as the publication of our initial study in January 2021.^2^ In order to reduce the risk of falsely negative diagnostic reports, the quality of HHM diagnostic testing must be improved so that the most common drivers of HHMs, such as *CHEK2* and *DDX41*, as well as highly penetrant HHMs, such as *ETV6, GATA2*, and *RUNX1*, are universally sequenced and analyzed.

## Data Availability

All data produced in the present work are contained in the manuscript

## Notes

### Competing Interest Statement

The authors have declared no competing interest.

