## Supplemental Figures for "Stagnation in quality of next-generation sequencing assays for the diagnosis of hereditary hematopoietic malignancies"

Contents:

S1. Supplementary Figure 1

S2. Supplementary Figure 2

S3. Supplementary Table 1

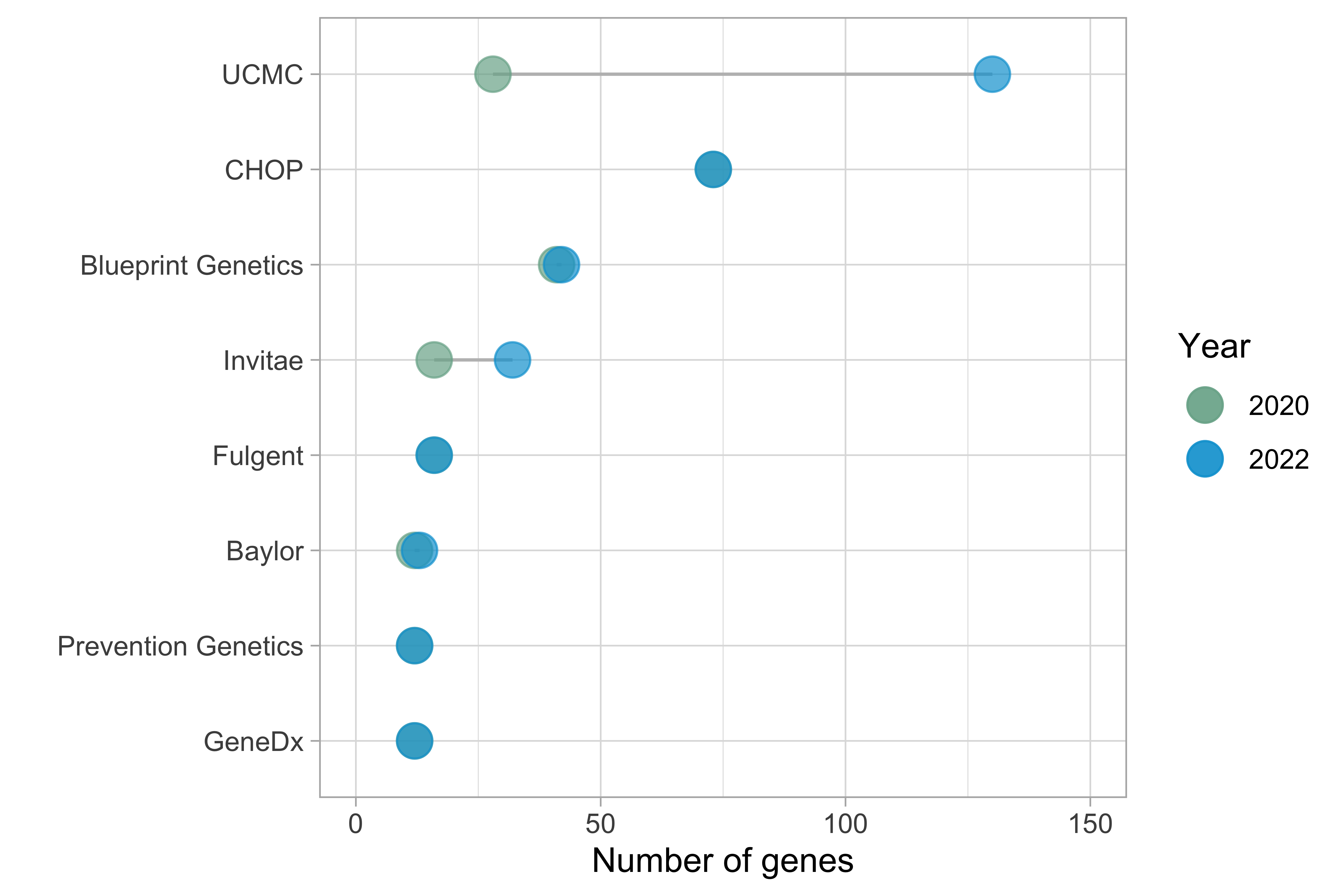

**Supplementary Figure 1.** Number of genes sequenced on each HHM panel in 2020 (green) and 2022 (blue). Abbreviations used: UCMC HMM, University of Chicago Medical Center Hereditary Myeloid Malignancy Panel ; CHOP, Children’s Hospital of Philadelphia.

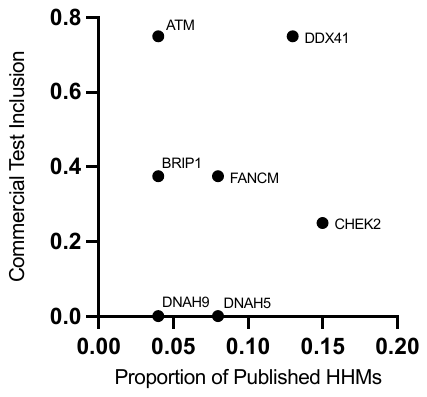

**Supplementary Figure 2.** Proportion of genes mutated in a case series of patients with HHMs (x axis, Yang *et al.*, *Blood*, 2022) versus proportion of these genes sequenced on commercially available HHM assays (y axis). Abbreviations used: HHM(s), hereditary hematopoietic malignancies.

**Supplementary Table 1 (separate excel file).** Online Mendelian Inheritance in Man phenotypes and numbers for reach gene sequenced on at least one HHM panel.
