## Supplemental Table 1 for "Stagnation in quality of next-generation sequencing assays for the diagnosis of hereditary hematopoietic malignancies"

| Gene | OMIM phenotype | OMIM Number |
|---|---|---|
| *ACD* | ?Dyskeratosis congenita, autosomal dominant 6, | 616553 |
| | ?Dyskeratosis congenita, autosomal recessive 7 | |
| *ADA2* | Sneddon syndrome | 182410 |
| | Vasculitis, autoinflammation, immunodeficiency, and hematologic defects syndrome | 615688 |
| *AK2* | Reticular dysgenesis | 267500 |
| *ALAS2* | Anemia, sideroblastic, 1 | 300751 |
| | Protoporphyria, erythropoietic, X-linked | 300752 |
| *ANKRD26* | Thrombocytopenia 2 | 188000 |
| *APC* | Adenoma, periampullary, somatic | 175100 |
| | Adenomatous polyposis coli | 175100 |
| | Brain tumor-polyposis syndrome 2 | 175100 |
| | Colorectal cancer, somatic | 114500 |
| | Desmoid disease, hereditary | 135290 |
| | Gardner syndrome | 175100 |
| | Gastric adenocarcinoma and proximal polyposis of the stomach | 619182 |
| | Gastric cancer, somatic | 613659 |
| | Hepatoblastoma, somatic | 11455 |
| *ATG2B* | NA | NA |
| *ATM* | Ataxia-telangiectasia | 208900 |
| | Lymphoma, B-cell non-Hodgkin, somatic | NA |
| | Lymphoma, mantle cell, somatic | NA |
| | T-cell prolymphocytic leukemia, somatic | NA |
| | {Breast cancer, susceptibility to} | 114480 |
| *ATR* | ?Cutaneous telangiectasia and cancer syndrome, familial | 614564 |
| | Seckel syndrome 1 | 210600 |
| *BLM* | Bloom syndrome | 210900 |
| *BRAF* | Adenocarcinoma of lung, somatic | 211980 |
| | Cardiofaciocutaneous syndrome | 115150 |
| | Colorectal cancer, somatic | 114500 |
| | LEOPARD syndrome 3 | 613707 |
| | Melanoma, malignant, somatic | 155600 |
| | Nonsmall cell lung cancer, somatic | 211980 |
| | Noonan syndrome 7 | 613706 |
| *BRCA1* | Fanconi anemia, complementation group S | 617883 |
| | {Breast-ovarian cancer, familial, 1} | 604370 |
| | {Pancreatic cancer, susceptibility to, 4} | 614320 |
| *BRCA2* | Fanconi anemia, complementation group D1 | 605724 |
| | Wilms tumor | 194070 |
| | {Breast cancer, male, susceptibility to} | 114480 |
| | {Breast-ovarian cancer, familial, 2} | 612555 |
| | {Glioblastoma 3} | 613029 |
| | {Medulloblastoma} | 155255 |
| | {Pancreatic cancer 2} | 613347 |
| | {Prostate cancer} | 176807 |
| *RIP1 (FANCJ* | Fanconi anemia, complementation group J | 609054 |
| | {Breast cancer, early-onset, susceptibility to} | 114480 |
| *CBL* | ?Juvenile myelomonocytic leukemia | 607785 |
| | Noonan syndrome-like disorder with or without juvenile myelomonocytic leukemia | 613563 |
| *CDKN2A* | {Melanoma and neural system tumor syndrome} | 155755 |
| | {Melanoma, cutaneous malignant, 2} | 155601 |
| | {Melanoma-pancreatic cancer syndrome} | 606719 |
| *CEBPA* | ?Leukemia, acute myeloid | 601626 |
| | Leukemia, acute myeloid, somatic | 601626 |
| *CHEK2* | Li-Fraumeni syndrome 2 | 609265 |
| | Osteosarcoma, somatic | 259500 |
| | {Breast cancer, susceptibility to} | 114480 |
| | {Colorectal cancer, susceptibility to} | 114500 |
| | {Prostate cancer, familial, susceptibility to} | 176807 |
| *CSF3R* | ?Neutrophilia, hereditary | 162830 |
| | Neutropenia, severe congenital, 7, autosomal recessive | 617014 |
| *CTC1* | Cerebroretinal microangiopathy with calcifications and cysts | 612199 |
| *DDX11* | Warsaw breakage syndrome | 613398 |
| *DDX41* | {Myeloproliferative/lymphoproliferative neoplasms, familial (multiple types), susceptibility to} | 616871 |
| *DKC1* | Dyskeratosis congenita, X-linked | 305000 |
| *DNAJC21* | Bone marrow failure syndrome 3 | 617052 |
| *FL1 / EFTUD* | Shwachman-Diamond syndrome 2 | 617941 |
| *ELANE* | Neutropenia, cyclic | 162800 |
| | Neutropenia, severe congenital 1, autosomal dominant | 202700 |
| *EPCAM* | Colorectal cancer, hereditary nonpolyposis, type 8 | 613244 |
| | Diarrhea 5, with tufting enteropathy, congenital | 613217 |
| *ERCC4* | Fanconi anemia, complementation group Q | 615272 |
| | Xeroderma pigmentosum, group F | 278760 |
| | Xeroderma pigmentosum, type F/Cockayne syndrome | 278760 |
| | XFE progeroid syndrome | 610965 |
| *ERCC6L2* | Bone marrow failure syndrome 2 | 615715 |
| *ESCO2* | Juberg-Hayward syndrome | 216100 |
| | Roberts-SC phocomelia syndrome | 268300 |
| *ETV6* | Leukemia, acute myeloid, somatic | 601626 |
| | Thrombocytopenia 5 | 616216 |
| *FANCA* | Fanconi anemia, complementation group A | 227650 |
| *FANCB* | Fanconi anemia, complementation group B | 300514 |
| *FANCC* | Fanconi anemia, complementation group C | 227645 |
| *FANCD2* | Fanconi anemia, complementation group D2 | 227646 |
| *FANCE* | Fanconi anemia, complementation group E | 600901 |
| *FANCF* | Fanconi anemia, complementation group F | 603467 |
| *FANCG* | Fanconi anemia, complementation group G | 614082 |
| *FANCI* | Fanconi anemia, complementation group I | 609053 |
| *FANCL* | Fanconi anemia, complementation group L | 614083 |
| *FANCM* | ?Premature ovarian failure 15 | 618096 |
| | Spermatogenic failure 28 | 618086 |
| *G6PC3* | Dursun syndrome | 612541 |
| | Neutropenia, severe congenital 4, autosomal recessive | 612541 |
| *GATA1* | Anemia, X-linked, with/without neutropenia and/or platelet abnormalities | 300835 |
| | Leukemia, megakaryoblastic, with or without Down syndrome, somatic | 190685 |
| | Thrombocytopenia with beta-thalassemia, X-linked | 314050 |
| | Thrombocytopenia, X-linked, with or without dyserythropoietic anemia | 300367 |
| *GATA2* | Emberger syndrome | 614038 |
| | Immunodeficiency 21 | 614172 |

| | | |
|---|---|---|
| [illegible] | {Leukemia, acute myeloid, susceptibility to} | 601626 |
| | {Myelodysplastic syndrome, susceptibility to} | 614286 |
| *GFI1* | ?Neutropenia, nonimmune chronic idiopathic, of adults | 607847 |
| | Neutropenia, severe congenital 2, autosomal dominant | 613107 |
| *GSKIP* | NA | NA |
| *HAVCR2* | T-cell lymphoma, subcutaneous panniculitis-like | 618398 |
| *HAX1* | Neutropenia, severe congenital 3, autosomal recessive | 610738 |
| *HRAS* | Bladder cancer, somatic | 109800 |
| | Congenital myopathy with excess of muscle spindles | 218040 |
| | Costello syndrome | 218040 |
| | Nevus sebaceous or woolly hair nevus, somatic | 162900 |
| | Schimmelpenning-Feuerstein-Mims syndrome, somatic mosaic | 163200 |
| | Spitz nevus or nevus spilus, somatic | 137550 |
| | Thyroid carcinoma, follicular, somatic | 188470 |
| *IKZF1* | Immunodeficiency, common variable, 13 | 616873 |
| *JAK2* | Erythrocytosis, somatic | 133100 |
| | Leukemia, acute myeloid, somatic | 601626 |
| | Myelofibrosis, somatic | 254450 |
| | Polycythemia vera, somatic | 263300 |
| | Thrombocythemia 3 | 614521 |
| | {Budd-Chiari syndrome, somatic} | 600880 |
| *KDM1A* | Cleft palate, psychomotor retardation, and distinctive facial features | 616728 |
| *KRAS* | Arteriovenous malformation of the brain, somatic | 108010 |
| | Bladder cancer, somatic | 109800 |
| | Breast cancer, somatic | 114480 |
| | Cardiofaciocutaneous syndrome 2 | 615278 |
| | Gastric cancer, somatic | 613659 |
| | Leukemia, acute myeloid, somatic | 601626 |
| | Lung cancer, somatic | 211980 |
| | Noonan syndrome 3 | 609942 |
| | Oculoectodermal syndrome, somatic | 600268 |
| | Pancreatic carcinoma, somatic | 260350 |
| | RAS-associated autoimmune leukoproliferative disorder | 614470 |
| | Schimmelpenning-Feuerstein-Mims syndrome, somatic mosaic | 163200 |
| *LIG4* | LIG4 syndrome | 606593 |
| | {Multiple myeloma, resistance to} | 254500 |
| *MAP2K1* | Cardiofaciocutaneous syndrome 3 | 615279 |
| | Melorheostosis, isolated, somatic mosaic | 155950 |
| *MAP2K2* | Cardiofaciocutaneous syndrome 4 | 615280 |
| *MBD4* | NA | NA |
| *MECOM* | Radioulnar synostosis with amegakaryocytic thrombocytopenia 2 | 616738 |
| *MLH1* | Colorectal cancer, hereditary nonpolyposis, type 2 | 609310 |
| | Mismatch repair cancer syndrome 1 | 276300 |
| | Muir-Torre syndrome | 158320 |
| *MPL* | Myelofibrosis with myeloid metaplasia, somatic | 254450 |
| | Thrombocythemia 2 | 601977 |
| | Thrombocytopenia, congenital amegakaryocytic | 604498 |
| *MSH2* | Colorectal cancer, hereditary nonpolyposis, type 1 | 120435 |
| | Mismatch repair cancer syndrome 2 | 619096 |
| | Muir-Torre syndrome | 158320 |
| *MSH6* | Colorectal cancer, hereditary nonpolyposis, type 5 | 614350 |
| | Mismatch repair cancer syndrome 3 | 619097 |
| | {Endometrial cancer, familial} | 608089 |
| *NAF1* | NA | NA |
| *NBN* | Aplastic anemia | 609135 |
| | Leukemia, acute lymphoblastic | 613065 |
| | Nijmegen breakage syndrome | 251260 |
| *NF1* | Leukemia, juvenile myelomonocytic | 607785 |
| | Neurofibromatosis, familial spinal | 162210 |
| | Neurofibromatosis, type 1 | 162200 |
| | Neurofibromatosis-Noonan syndrome | 601321 |
| | Watson syndrome | 193520 |
| *NF2* | Meningioma, NF2-related, somatic | 607174 |
| | Neurofibromatosis, type 2 | 101000 |
| | Schwannomatosis, somatic | 162091 |
| *NHP2* | Dyskeratosis congenita, autosomal recessive 2 | 613987 |
| *NHEJ1* | Severe combined immunodeficiency with microcephaly, growth retardation, and sensitivity to ionizing radiation | 611291 |
| *NOP10* | Dyskeratosis congenita, autosomal recessive 1 | 224230 |
| *NPM1* | Leukemia, acute myeloid, somatic | 601626 |
| *NRAS* | ?RAS-associated autoimmune lymphoproliferative syndrome type IV, somatic | 614470 |
| | Colorectal cancer, somatic | 114500 |
| | Epidermal nevus, somatic | 162900 |
| | Melanocytic nevus syndrome, congenital, somatic | 137550 |
| | Neurocutaneous melanosis, somatic | 249400 |
| | Noonan syndrome 6 | 613224 |
| | Schimmelpenning-Feuerstein-Mims syndrome, somatic mosaic | 163200 |
| | Thyroid carcinoma, follicular, somatic | 188470 |
| *PALB2* | Fanconi anemia, complementation group N | 610832 |
| | {Breast cancer, susceptibility to} | 114480 |
| | {Pancreatic cancer, susceptibility to, 3} | 613348 |
| *PARN* | Dyskeratosis congenita, autosomal recessive 6 | 616353 |
| | Pulmonary fibrosis and/or bone marrow failure, telomere-related, 4 | 616371 |
| *PAX5* | {Leukemia, acute lymphoblastic, susceptibility to, 3} | 615545 |
| *PMS2* | Colorectal cancer, hereditary nonpolyposis, type 4 | 614337 |
| | Mismatch repair cancer syndrome 4 | 619101 |
| *POT1* | {Glioma susceptibility 9} | 616568 |
| | {Melanoma, cutaneous malignant, susceptibility to, 10} | 615848 |
| *PRF1* | Aplastic anemia | 609135 |
| | Hemophagocytic lymphohistiocytosis, familial, 2 | 603553 |
| | Lymphoma, non-Hodgkin | 605027 |
| *PTPN11* | LEOPARD syndrome 1 | 151100 |
| | Leukemia, juvenile myelomonocytic, somatic | 607785 |
| | Metachondromatosis | 156250 |
| | Noonan syndrome 1 | 163950 |
| *RAD51* | Fanconi anemia, complementation group R | 617244 |
| | Mirror movements 2 | 614508 |
| | {Breast cancer, susceptibility to} | 114480 |
| *RAD51C* | Fanconi anemia, complementation group O | 613390 |
| | {Breast-ovarian cancer, familial, susceptibility to, 3} | 613399 |
| | Cardiomyopathy, dilated, 1NN | 615916 |

| | | |
|---|---|---|
| *RAF1* | LEOPARD syndrome 2 | 611554 |
| | Noonan syndrome 5 | 611553 |
| *RBBP6* | NA | NA |
| *RBM8A* | Thrombocytopenia-absent radius syndrome | 274000 |
| *RECQL* | NA | NA |
| *RECQL4* | Baller-Gerold syndrome | 218600 |
| | RAPADILINO syndrome | 266280 |
| | Rothmund-Thomson syndrome, type 2 | 268400 |
| *RMRP* | Anauxetic dysplasia 1 | 607095 |
| | Cartilage-hair hypoplasia | 250250 |
| | Metaphyseal dysplasia without hypotrichosis | 250460 |
| *RPL11* | Diamond-Blackfan anemia 7 | 612562 |
| *RPL15* | ?Diamond-Blackfan anemia 12 | 615550 |
| *RPL18* | ?Diamond-Blackfan anemia 18 | 618310 |
| *RPL23* | NA | NA |
| *RPL26* | ?Diamond-Blackfan anemia 11 | 614900 |
| *RPL27* | ?Diamond-Blackfan anemia 16 | 617408 |
| *RPL31* | NA | NA |
| *RPL35* | ?Diamond-Blackfan anemia 19 | 618312 |
| *RPL35A* | Diamond-Blackfan anemia 5 | 612528 |
| *RPL36* | NA | NA |
| *RPL5* | Diamond-Blackfan anemia 6 | 612561 |
| *RPS10* | Diamond-Blackfan anemia 9 | 613308 |
| *RPS17* | Diamond-Blackfan anemia 4 | 612527 |
| *RPS19* | Diamond-Blackfan anemia 1 | 105650 |
| *RPS24* | Diamond-blackfan anemia 3 | 610629 |
| *RPS26* | Diamond-Blackfan anemia 10 | 613309 |
| *RPS27* | ?Diamond-Blackfan anemia 17 | 617409 |
| *RPS28* | Diamond Blackfan anemia 15 with mandibulofacial dysostosis | 606164 |
| *RPS29* | Diamond-Blackfan anemia 13 | 615909 |
| *RPS7* | Diamond-Blackfan anemia 8 | 612563 |
| *RIT1* | Noonan syndrome 8 | 615355 |
| *RTEL1* | Dyskeratosis congenita, autosomal dominant 4 | 615190 |
| | Dyskeratosis congenita, autosomal recessive 5 | 615190 |
| | Pulmonary fibrosis and/or bone marrow failure, telomere-related, 3 | 616373 |
| *RUNX1* | Leukemia, acute myeloid | 601626 |
| | Platelet disorder, familial, with associated myeloid malignancy | 601399 |
| *SAMD9* | MIRAGE syndrome | 617053 |
| | Monosomy 7 myelodysplasia and leukemia syndrome 2 | 619041 |
| | Tumoral calcinosis, familial, normophosphatemic | 610455 |
| *SAMD9L* | Ataxia-pancytopenia syndrome | 159550 |
| | Monosomy 7 myelodysplasia and leukemia syndrome 1 | 252270 |
| | Spinocerebellar ataxia 49 | 619806 |
| *SBDS* | Shwachman-Diamond syndrome | 260400 |
| | {Aplastic anemia, susceptibility to} | 609135 |
| *SH2B3* | Erythrocytosis, somatic | 133100 |
| | Myelofibrosis, somatic | 254450 |
| | Thrombocythemia, somatic | 187950 |
| *SLX4* | Fanconi anemia, complementation group P | 613951 |
| *SOS1* | ?Fibromatosis, gingival, 1 | 135300 |
| | Noonan syndrome 4 | 610733 |
| *SRP54* | Neutropenia, severe congenital, 8, autosomal dominant | 618752 |
| *SRP72* | Bone marrow failure syndrome 1 | 614675 |
| *TERC* | Dyskeratosis congenita, autosomal dominant 1 | 127550 |
| | {Aplastic anemia} | 614743 |
| | {Pulmonary fibrosis, idiopathic, susceptibility to} | 614743 |
| *TERT* | Dyskeratosis congenita, autosomal dominant 2 | 613989 |
| | Dyskeratosis congenita, autosomal recessive 4 | 613989 |
| | Pulmonary fibrosis and/or bone marrow failure, telomere-related, 1 | 614742 |
| | {Leukemia, acute myeloid} | 601626 |
| | {Melanoma, cutaneous malignant, 9} | 615134 |
| *TET2* | Immunodeficiency 75 | 619126 |
| | Myelodysplastic syndrome, somatic | 614286 |
| *TINF2* | Dyskeratosis congenita, autosomal dominant 3 | 613990 |
| | Revesz syndrome | 268130 |
| *TP53* | Bone marrow failure syndrome 5 | 618165 |
| | Breast cancer, somatic | 114480 |
| | Hepatocellular carcinoma, somatic | 114550 |
| | Li-Fraumeni syndrome | 151623 |
| | Nasopharyngeal carcinoma, somatic | 607107 |
| | Pancreatic cancer, somatic | 260350 |
| | {Adrenocortical carcinoma, pediatric} | 202300 |
| | {Basal cell carcinoma 7} | 614740 |
| | {Choroid plexus papilloma} | 260500 |
| | {Colorectal cancer} | 114500 |
| | {Glioma susceptibility 1} | 137800 |
| | {Osteosarcoma} | 259500 |
| *UBE2T* | Fanconi anemia, complementation group T | 616435 |
| *USB1* | Poikiloderma with neutropenia | 604173 |
| *VPS45* | Neutropenia, severe congenital, 5, autosomal recessive | 615285 |
| *WAS* | Wiskott-Aldrich syndrome | 301000 |
| *WIPF1* | Wiskott-Aldrich syndrome 2 | 614493 |
| *WRAP53* | Dyskeratosis congenita, autosomal recessive 3 | 613988 |
| *XRCC2* | ?Fanconi anemia, complementation group U | 617247 |
| | ?Premature ovarian failure 17 | 619146 |
| | Spermatogenic failure 50 | 619145 |
